# Peripheral Blood NADP(H) Redox Metabolism in Alzheimer’s Disease and Mild Cognitive Impairment: A Systematic Review and Meta-analysis Where Feasible

**DOI:** 10.64898/2026.11.09.26367672

**Authors:** Dongsheng Zhong, Dongquan He, Jia Chen, Chenkai Yanyang

## Abstract

**Background:** The reduced and oxidized forms of nicotinamide adenine dinucleotide phosphate (NADPH/NADP□) connect cellular reducing power with the glutathione and thioredoxin systems. Peripheral alterations in this network may reflect systemic redox dysregulation in Alzheimer’s disease (AD) and mild cognitive impairment (MCI), but the evidence has not been quantitatively synthesized.

**Objective:** To systematically evaluate peripheral-blood NADP(H) redox metabolism and related enzymes in AD and MCI and to perform meta-analyses when the available data were sufficiently comparable.

**Methods:** PubMed, Embase, and Web of Science were searched from inception to 6 November 2026. Eligible observational studies compared patients with AD or MCI with cognitively unimpaired controls and reported NADPH, NADP□, the NADPH/NADP□ ratio, or NADPH-related enzymes, including glutathione reductase (GR), glucose-6-phosphate dehydrogenase (G6PD), and thioredoxin reductase (TRXR). Continuous outcomes were converted to Hedges g. Random-effects models used restricted maximum likelihood estimation and modified Hartung–Knapp confidence intervals.

**Results:** Of 2,686 records identified, 1,781 remained after deduplication and 13 studies were included. No study directly reported peripheral NADPH, NADP□, or the NADPH/NADP□ ratio. The pooled effect for erythrocyte GR in AD was g=−1.05 (95% CI −2.73 to 0.63; I^2^=95.0%; five studies); for erythrocyte GR in MCI, g=−0.15 (95% CI −8.68 to 8.38; I^2^=95.0%; two studies); for whole-blood/plasma GR in AD, g=−0.03 (95% CI −4.37 to 4.30; I^2^=97.1%; three studies); and for erythrocyte G6PD in AD, g=−0.66 (95% CI −17.36 to 16.03; I^2^=96.4%; two studies). Leave-one-out analyses did not materially alter the interpretation of the erythrocyte GR synthesis.

**Conclusions:** AD and MCI may be accompanied by remodeling of peripheral NADP(H)-related enzyme networks, but the direction and magnitude of effects are inconsistent across studies. Direct measurement of NADPH, NADP□, and their ratio remains the most important evidence gap. Future prospective studies should standardize specimen type, pre-analytical handling, and analytical platforms and should account for disease stage, treatment exposure, and metabolic comorbidity.

## 1. Introduction

Alzheimer’s disease (AD) is a progressive neurodegenerative disorder characterized clinically by cognitive and functional decline. Mild cognitive impairment (MCI) occupies an intermediate clinical space between healthy aging and dementia and is a key stage for identifying early biological changes. Alongside amyloid-β deposition, tau pathology, synaptic dysfunction, and neuroinflammation, disruption of redox homeostasis is increasingly recognized as an integral component of AD pathobiology [15–17,27]. Peripheral blood captures systemic metabolic, inflammatory, and antioxidant responses and therefore provides an accessible window into disease-associated biology [24–26].

The NADP(H) system is a central hub of reductive metabolism. NADPH supplies reducing equivalents to glutathione reductase, thioredoxin reductase, and multiple antioxidant and biosynthetic reactions. Glucose-6-phosphate dehydrogenase (G6PD), the rate-controlling enzyme of the oxidative pentose phosphate pathway, contributes substantially to cytosolic NADPH regeneration, particularly in erythrocytes [21–23]. The NADPH/NADP□ ratio is determined by production, consumption, and compartment-specific transport and is more directly related to cellular reducing capacity than any single antioxidant enzyme [21,22,33,34].

Case–control studies have reported altered peripheral GR, G6PD, and TRXR activity in AD and MCI, but their findings vary in direction and magnitude [1–13]. These studies also differ in specimen type, assay platform, disease severity, treatment status, and reporting units. Importantly, direct quantification of NADPH and NADP□ addresses a different biological question from measurement of enzymes that generate or consume NADPH. We therefore performed a systematic review centered specifically on peripheral NADP(H) metabolism, distinguished direct redox-pair measurements from pathway-proximal enzyme markers, and conducted stratified meta-analyses where the evidence was sufficiently comparable.

## 2. Methods

### 2.1 Study design and registration

This systematic review was designed, conducted, and reported in accordance with the PRISMA 2020 statement [14]. The review protocol is intended for registration in PROSPERO (registration number: CRD420261566620). General methodological guidance was also informed by the Cochrane Handbook [45].

### 2.2 Search strategy

PubMed, Embase, and Web of Science were searched from database inception to 6 November 2026. The strategy combined terms for the disease or cognitive stage (e.g., “Alzheimer disease,” “dementia,” and “mild cognitive impairment”) with terms for NADP(H) metabolism (e.g., “NADPH,” “NADP,” “NADPH/NADP ratio,” “glutathione reductase,” “glucose-6-phosphate dehydrogenase,” and “thioredoxin reductase”). No language restriction was applied. Reference lists of included studies and relevant reviews were also screened. Full database-specific strategies are provided in the Supplementary Methods.

### 2.3 Eligibility criteria

Studies were eligible if they: (1) included clinically diagnosed AD or MCI and a cognitively unimpaired or healthy control group; (2) analyzed peripheral blood, whole blood, plasma, serum, erythrocytes, or peripheral blood cells; (3) reported NADPH, NADP□, the NADPH/NADP□ ratio, or an explicitly NADPH-dependent pathway marker such as GR, G6PD/G6PDH, or TRXR; (4) used an observational design or provided extractable baseline data from an intervention study; and (5) reported quantitative between-group data or an interpretable direction of effect. Animal, cell-culture, and brain-only studies; studies without an eligible comparator; reports restricted to oxidative-stress markers without a direct NADP(H) pathway relationship; reviews, comments, case reports, and duplicate publications were excluded. For overlapping cohorts, the most informative report was retained.

### 2.4 Study selection and data extraction

After deduplication, titles and abstracts were screened, followed by full-text assessment. Extracted variables included author, year, design, diagnostic group, sample size, specimen matrix, assay method, measurement unit, mean, and dispersion. Standard errors were converted to standard deviations using SD=SEM×√n. When an article reported multiple AD subgroups that required a single overall comparison, means and variances were combined using sample-size-weighted formulas. Graph-only data without a reliably extractable variance were retained for narrative synthesis. Studies with conflicting values between the main text and tables were excluded from the primary quantitative analysis and examined in scenario analyses. Conversions and distributional approximations followed established methodological guidance where applicable [43,44].

### 2.5 Outcomes and statistical analysis

The prespecified primary outcomes were peripheral NADPH, NADP□, and the NADPH/NADP□ ratio. Secondary outcomes were NADPH-related enzymes, including GR, G6PD/G6PDH, and TRXR. To limit biological non-comparability, analyses were stratified by disease stage, specimen matrix, and analyte. Continuous outcomes were standardized as Hedges g, defined as the patient group minus the control group; negative values therefore indicate lower activity or concentration in patients. Random-effects models used restricted maximum likelihood estimation of τ^2^ and modified Hartung–Knapp 95% confidence intervals [38–42]. Heterogeneity was described using Cochran Q and I^2^ [40]. Leave-one-out analyses were conducted for the principal synthesis, and scenario analyses were used for internally conflicting data. Funnel plots and Egger tests were not performed because every synthesis included fewer than 10 studies. Analyses were conducted in R version 4.6.1. Two-sided P<0.05 was considered statistically significant.

## 3. Results

### 3.1 Study selection

The searches identified 2,686 records: 819 from Embase, 578 from PubMed, and 1,289 from Web of Science. After removal of 905 duplicates, 1,781 records underwent title and abstract screening, of which 1,763 were excluded. Eighteen reports were sought for retrieval; four did not yield a separate full-text report suitable for independent assessment. Fourteen full texts were assessed, one was excluded because the reported indicator was ineligible or the retrieved article did not match the indexed title, and 13 studies were included (Figure 1).

**Figure 1.**
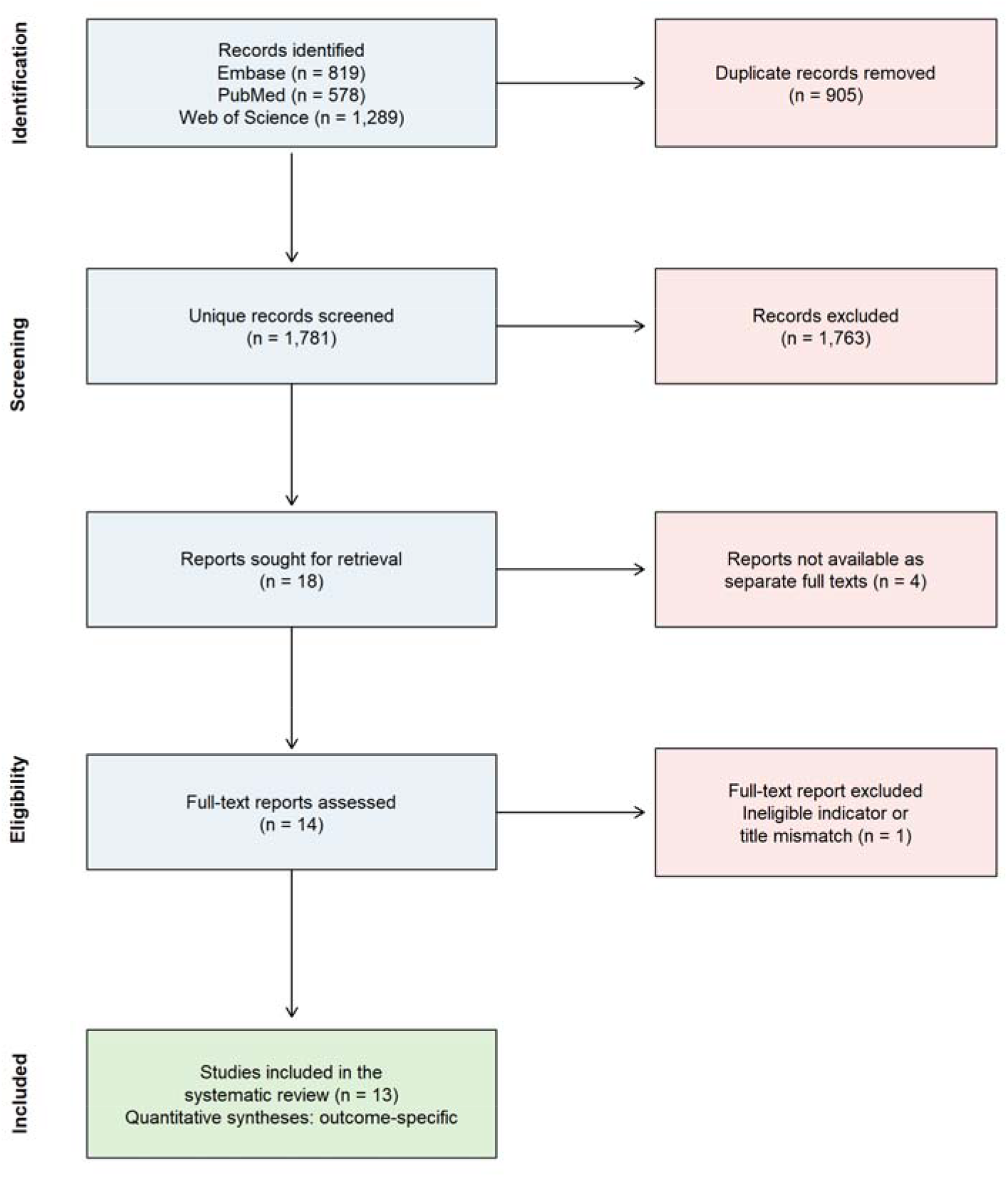
PRISMA 2020 flow diagram of study selection.

### 3.2 Study characteristics and the direct-evidence gap

The 13 studies were published between 1996 and 2023. Study groups included AD, MCI, and, in some reports, vascular dementia as a differential-diagnosis comparator. Specimens included erythrocytes, whole blood, plasma, serum, lymphocytes, and other peripheral blood cells. GR was the most frequently measured enzyme, followed by G6PD/G6PDH and TRXR (Table 1). None of the included studies directly reported peripheral NADPH, NADP□, or the NADPH/NADP□ ratio. Consequently, no quantitative synthesis was possible for the prespecified primary redox-pair outcomes, and the available meta-analyses addressed pathway-related enzymes. The analytical disposition of all prespecified outcomes is summarized in Table 5.

**Table 1.** Characteristics of included studies.

| **Study** | **Sample size** | **Matrix** | **Marker** | **Notes** |
|---|---|---|---|---|
| Park 2023 | AD 20; control 20 | Plasma | GR concentration | ELISA; cross-sectional |
| Sarhat 2019 | AD 54; control 54 | Serum | G6PD | Conflicting table/text data |
| Martínez de Toda 2019 | AD 20; MCI 20; control 30 | Peripheral blood cells | GR | Graph only; variance unavailable |
| Yuan 2016 | MCI 138; control 138 | Erythrocytes | GR | Case–control |
| Krishnan 2014 | AD 30; VaD 35; control 40 | Plasma/erythrocytes | GR, G6PD, TR | Multiple markers |
| Kosenko 2012 | AD 12; non-AD 13; older control 14; young control 14 | Erythrocytes | GR, G6PD | Age-matched control used |
| Klugman 2012 | Untreated AD 25; treated AD 43; control 34 | Whole blood | GR | Primary analysis used untreated AD |
| Torres 2011 | AD 29; MCI 33; control 26 | Erythrocytes | GR | Shared control group |
| Casado 2008 | AD 50; VaD 50; control 50 | Erythrocytes | GR | Case–control |
| Calabrese 2006 | AD 18; control 18 (figure subset 9/group) | Lymphocytes/plasma | TRXR | Graph only; brain tissue not pooled |
| Zafrilla 2006 | Mild/moderate AD 36; severe AD 30; control 27 | Whole blood | GR | AD subgroups combined |
| Liu 2005 | Men: AD 14/control 10; women: AD 19/control 10 | Erythrocytes | GR | Graph only |
| Ceballos-Picot 1996 | AD 40; control 34 | Erythrocytes | GR | Case–control |

### 3.3 Erythrocyte GR in AD

Five studies contributed to the meta-analysis of erythrocyte GR in AD. Although the pooled estimate was directed toward lower GR activity in AD, its confidence interval crossed the null (Hedges g=−1.05, 95% CI −2.73 to 0.63; τ^2^=1.73; I^2^=95.0%; Q=80.10, P<0.001; Figure 2).

**Figure 2.**
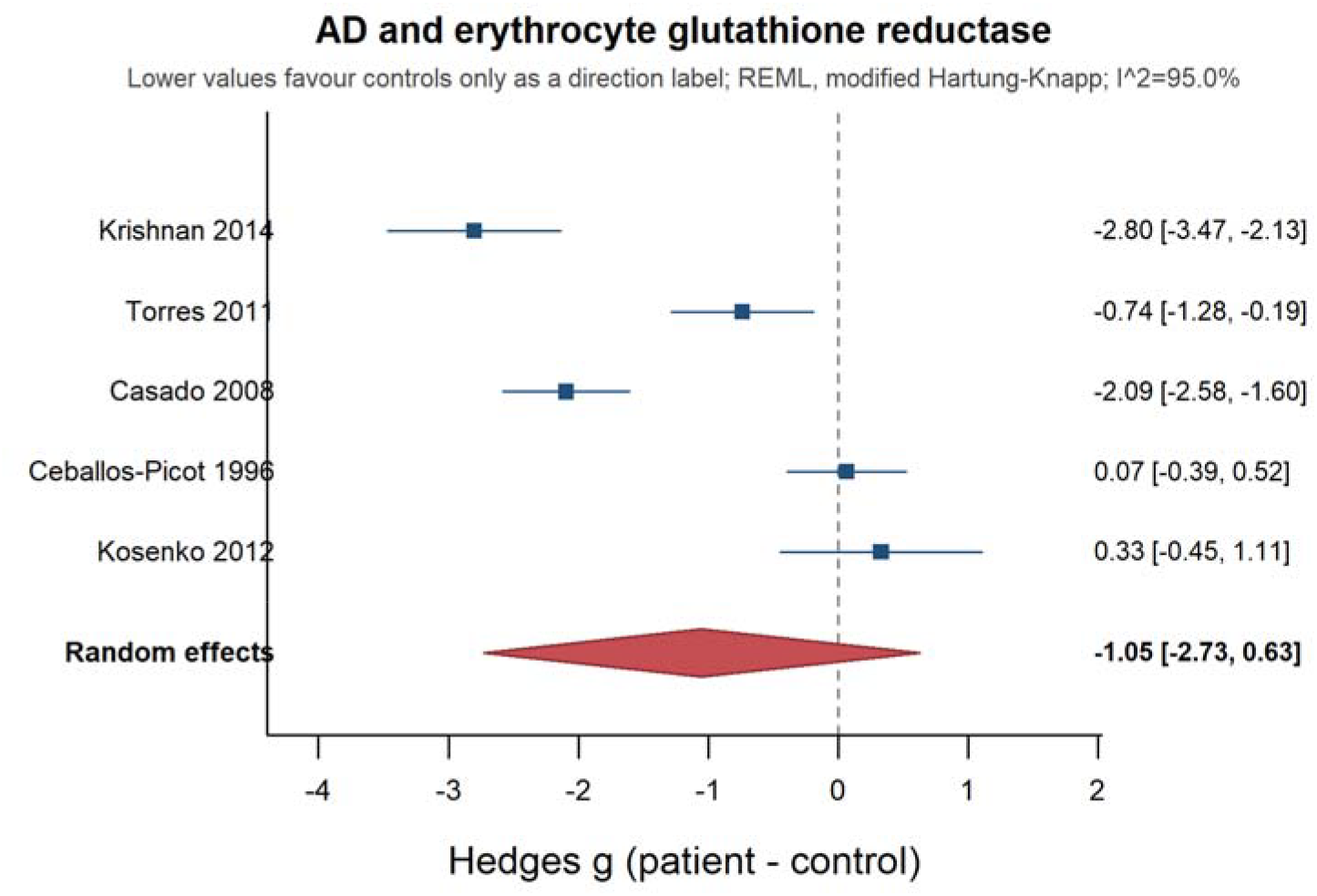
Random-effects meta-analysis of erythrocyte GR activity in AD. Squares indicate study-specific Hedges g, horizontal lines indicate 95% confidence intervals, and the diamond indicates the REML random-effects estimate. Negative values indicate lower activity in patients.

Individual study estimates were discordant: several showed marked reductions, whereas others were close to the null or modestly positive. Summary estimates for all quantitative syntheses are presented in Table 2.

**Table 2.**
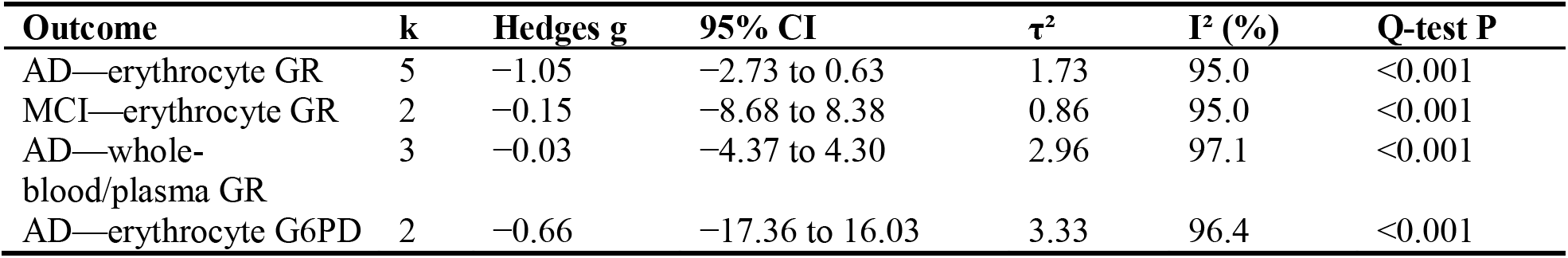
Main meta-analysis results.

### 3.4 Erythrocyte GR in MCI

Two studies reported erythrocyte GR in MCI. Their effects were in opposite directions, yielding an imprecise pooled estimate close to the null (Hedges g=−0.15, 95% CI −8.68 to 8.38; τ^2^=0.86; I^2^=95.0%; Q=20.06, P<0.001; Figure 3).

**Figure 3.**
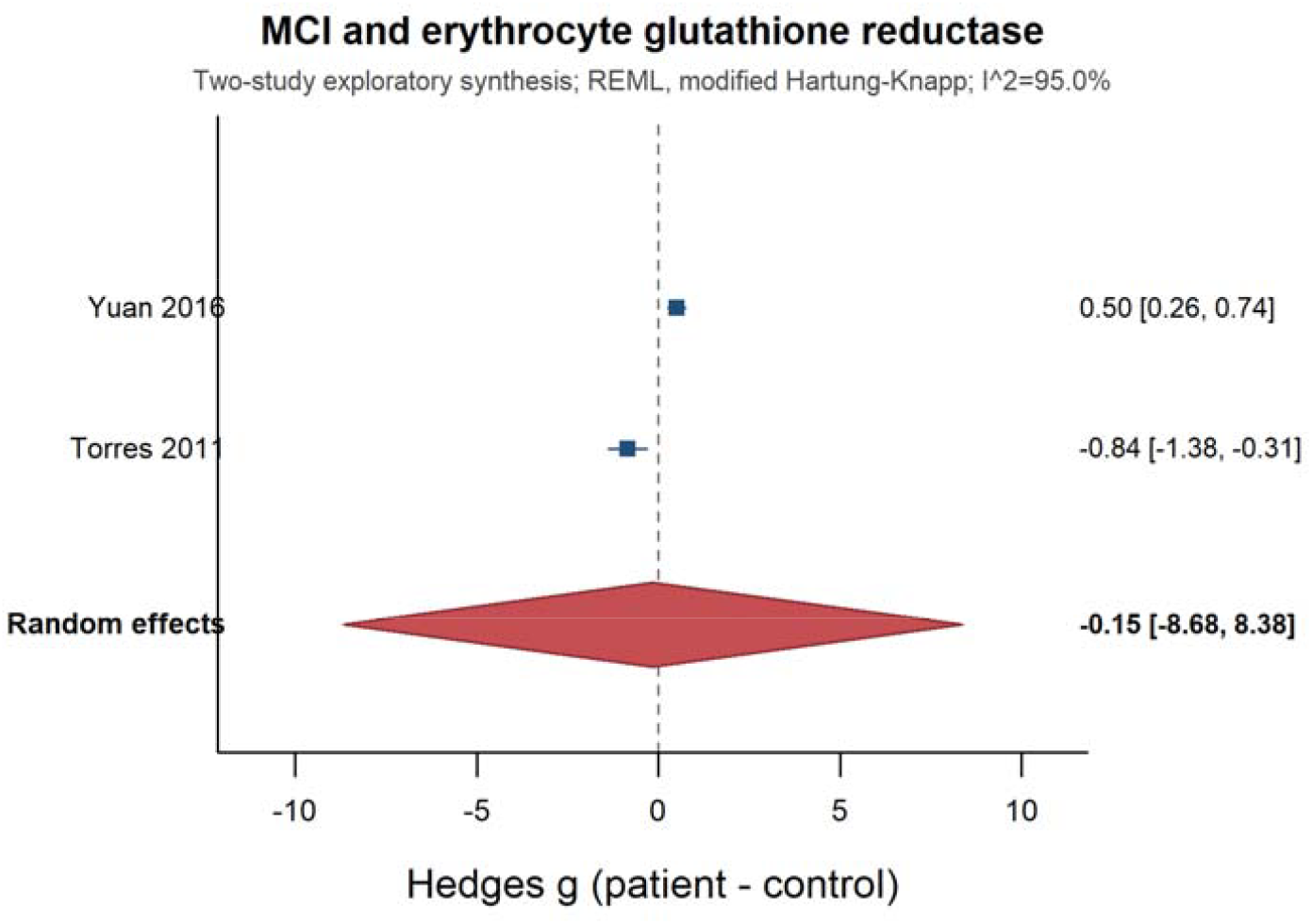
Exploratory random-effects meta-analysis of erythrocyte GR activity in MCI.

### 3.5 Whole-blood or plasma GR in AD

Three studies reported whole-blood or plasma GR in AD. The pooled estimate was close to zero (Hedges g=−0.03, 95% CI −4.37 to 4.30; τ^2^=2.96; I^2^=97.1%; Q=69.13, P<0.001; Figure 4).

**Figure 4.**
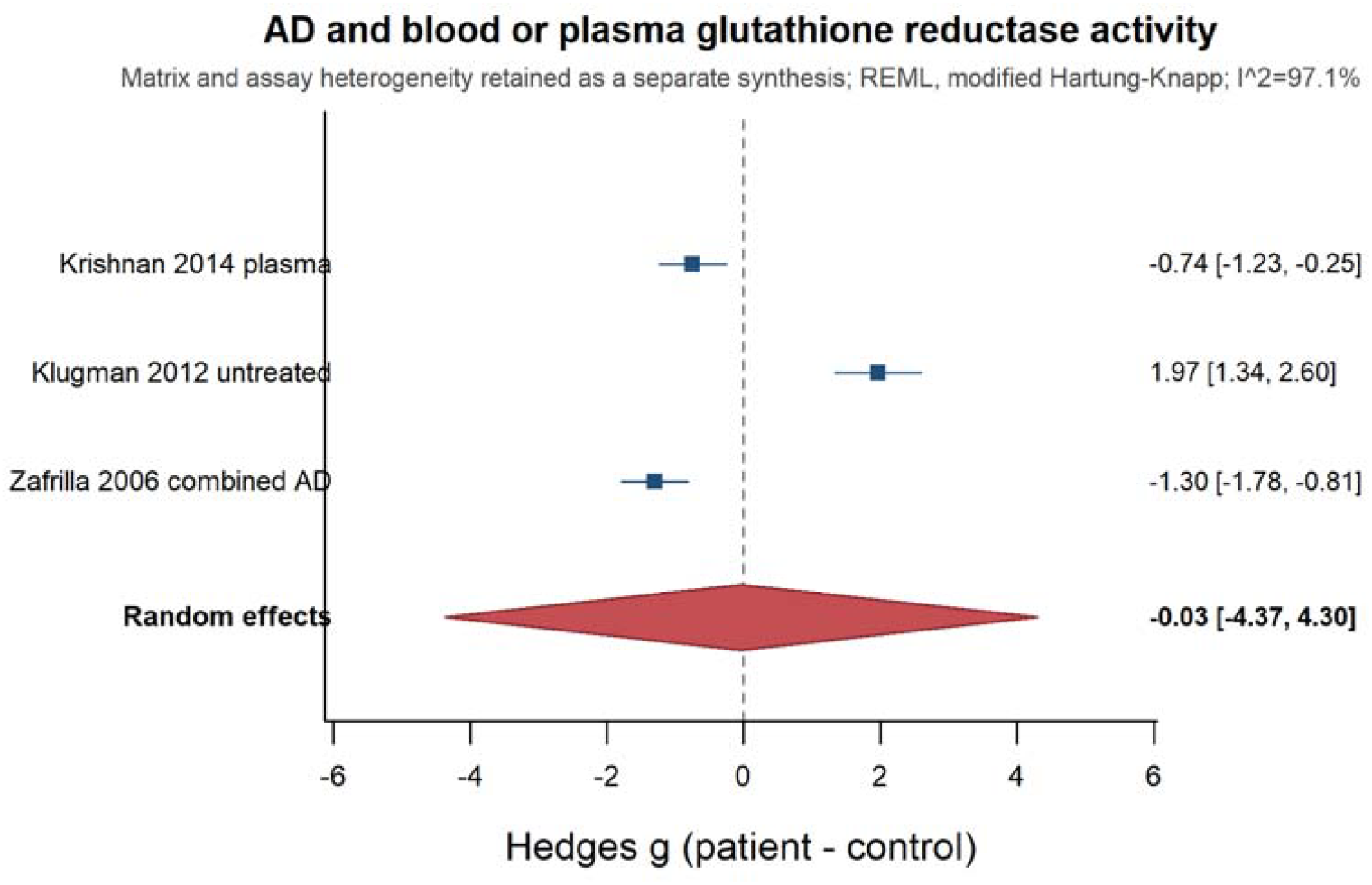
Exploratory random-effects meta-analysis of whole-blood or plasma GR activity in AD.

The component studies included lower plasma GR, higher whole-blood GR in untreated AD, and differing activities across disease-severity groups.

### 3.6 Erythrocyte G6PD in AD

The primary analysis included two studies with internally consistent and extractable data. The pooled estimate was directed toward lower G6PD activity in AD but was highly imprecise (Hedges g=−0.66, 95% CI −17.36 to 16.03; τ^2^=3.33; I^2^=96.4%; Q=27.56, P<0.001; Figure 5).

**Figure 5.**
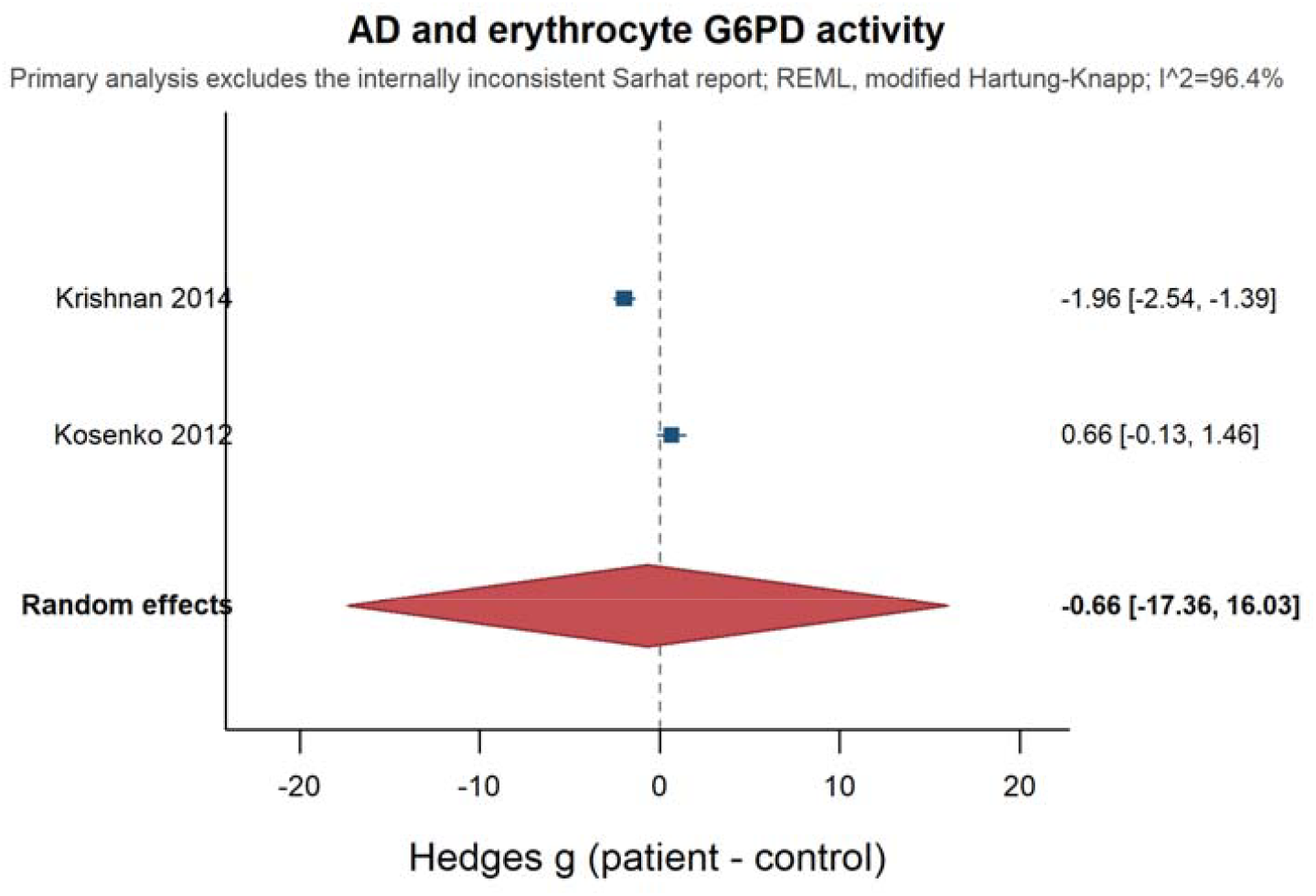
Random-effects meta-analysis of erythrocyte G6PD activity in AD; the primary analysis excluded the internally inconsistent Sarhat report.

The report by Sarhat et al. contained inconsistent dispersion values in the table and main text and was therefore excluded from the primary analysis [2].

### 3.7 Other NADP(H)-related markers

TRXR and several GR outcomes were available only as figures without complete means and variances. Narrative findings suggested perturbation of the peripheral thiol-reduction system in AD, but the small number of studies and differences in matrix and measurement scale precluded cross-analyte pooling.

### 3.8 Sensitivity analyses

After sequential omission of each erythrocyte GR study in AD, pooled Hedges g values ranged from −1.38 to −0.62; all 95% confidence intervals crossed zero and I^2^ remained between 93.8% and 96.2% (Table 3; Figure 6). Thus, the overall interpretation was not driven by a single study. In scenario analyses of G6PD, adding the tabulated Sarhat data yielded g=−11.66 (95% CI −59.48 to 36.16; I^2^=99.1%), whereas adding the values described in the main text yielded g=−2.01 (95% CI −8.68 to 4.66; I^2^=97.9%). Neither scenario produced a precise or consistent pooled effect. The G6PD scenario analyses are detailed in Table 4.

**Table 3.** Leave-one-out sensitivity analysis for erythrocyte GR in AD.

| Omitted study | k | Pooled Hedges g | 95% CI | I<sup>2</sup> (%) |
|---|---|---|---|---|
| Krishnan 2014 | 4 | −0.62 | −2.36 to 1.11 | 93.8 |
| Torres 2011 | 4 | −1.13 | −3.60 to 1.34 | 96.2 |
| Casado 2008 | 4 | −0.78 | −3.03 to 1.46 | 94.6 |
| Ceballos-Picot 1996 | 4 | −1.34 | −3.54 to 0.87 | 93.9 |
| Kosenko 2012 | 4 | −1.38 | −3.44 to 0.68 | 95.5 |

**Table 4.**
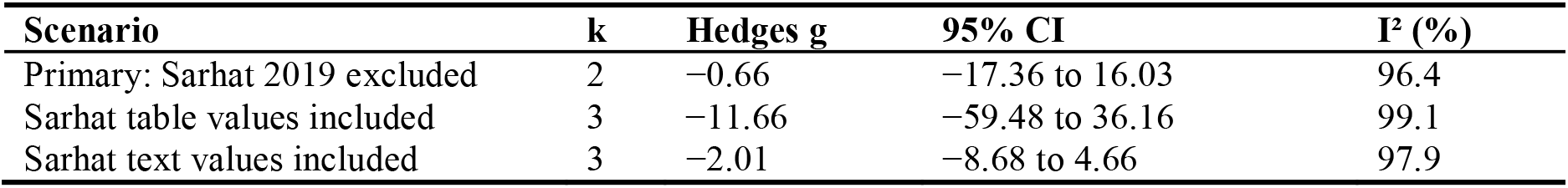
Scenario sensitivity analyses for erythrocyte G6PD in AD.

| Scenario | k | Hedges g | 95% CI | I<sup>2</sup> (%) |
|---|---|---|---|---|
| Primary: Sarhat 2019 excluded | 2 | −0.66 | −17.36 to 16.03 | 96.4 |
| Sarhat table values included | 3 | −11.66 | −59.48 to 36.16 | 99.1 |
| Sarhat text values included | 3 | −2.01 | −8.68 to 4.66 | 97.9 |

**Table 5.** Poolability and analytical handling of outcomes.

| **Outcome** | **Available studies** | **Evidence status** | **Analytical handling** |
|---|---|---|---|
| Direct NADPH, NADP□, or NADPH/NADP□ ratio | 0 | No direct data | No meta-analysis |
| AD erythrocyte GR | 5 | Comparable matrix and marker | Principal quantitative synthesis |
| MCI erythrocyte GR | 2 | Only two studies | Exploratory synthesis |
| AD whole-blood/plasma GR | 3 | Matrix and assay heterogeneity | Separate exploratory synthesis |
| AD erythrocyte G6PD | 2 | Internal conflict in Sarhat report | Excluded from primary; scenario analyses |
| TRXR and graph-only GR | Several | Mean/variance not reliably extractable | Narrative synthesis |

**Figure 6.**
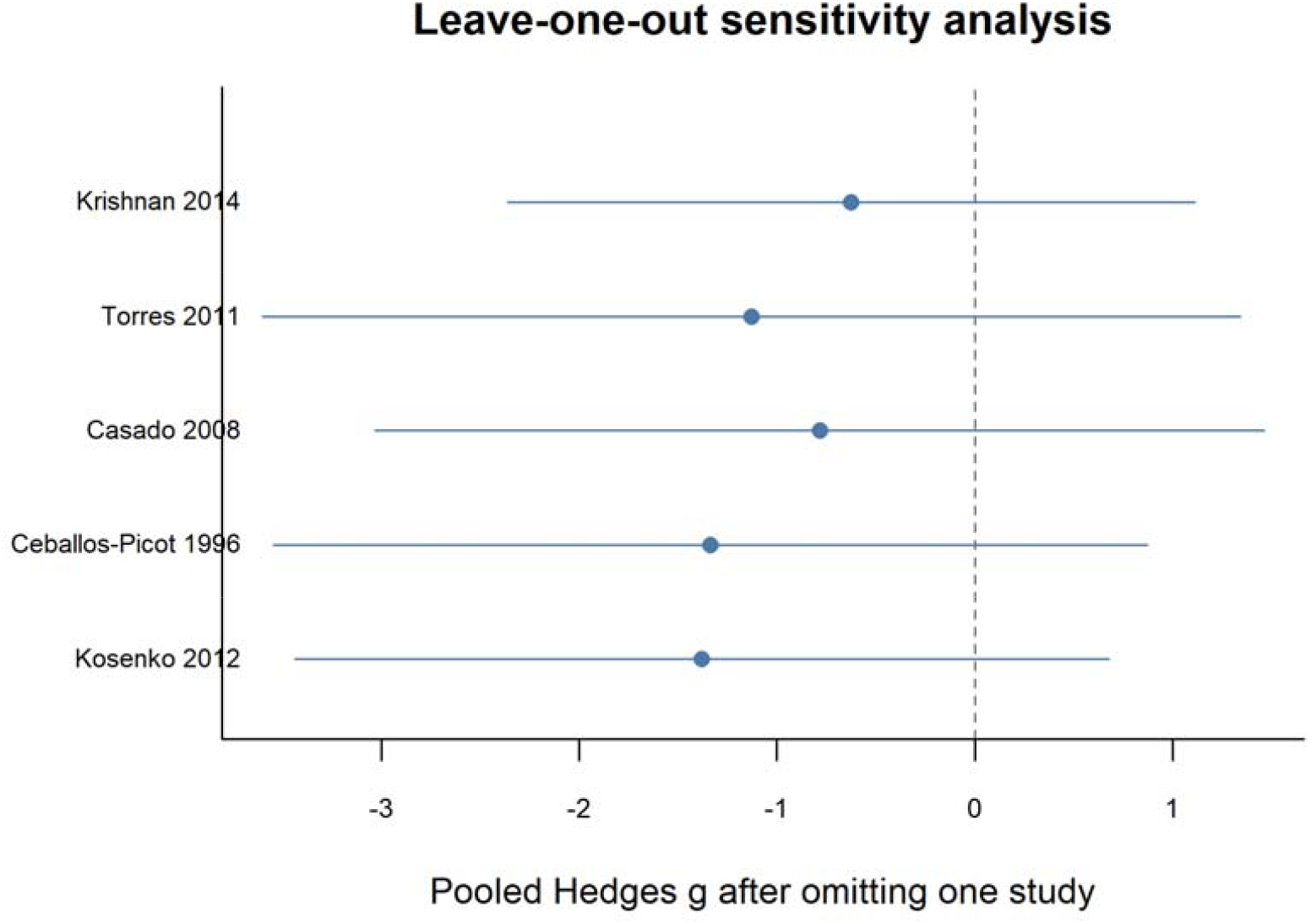
Leave-one-out sensitivity analysis of erythrocyte GR in AD. Points and horizontal lines indicate pooled Hedges g and 95% confidence intervals after omission of the specified study.

## 4. Discussion

### 4.2 Principal findings

This systematic review evaluated peripheral redox biology through the specific lens of NADP(H) metabolism. Its clearest finding is that no eligible study directly measured peripheral NADPH, NADP□, or the NADPH/NADP□ ratio. The quantitative evidence instead concerns enzymes that generate or use NADPH. Across these enzyme-based syntheses, pooled confidence intervals crossed the null and between-study heterogeneity was substantial. The current literature therefore supports the presence of peripheral redox remodeling in AD and MCI, but not a reproducible unidirectional change in any single NADP(H)-related enzyme.

### 4.3 Biological interpretation of the NADP(H) network

GR uses NADPH to reduce oxidized glutathione, G6PD replenishes cytosolic NADPH through the pentose phosphate pathway, and TRXR transfers NADPH-derived reducing equivalents to the thioredoxin system [20–23,29,32,37]. Lower GR may indicate impaired glutathione recycling; altered G6PD may reflect deficient NADPH supply or compensatory pathway activation; and TRXR changes may affect protein-thiol homeostasis. Nevertheless, enzyme abundance or activity is not equivalent to the NADPH/NADP□ ratio. Cofactor availability, substrate load, erythrocyte age distribution, oxidative demand, and compensatory regulation can generate opposite enzyme responses under similar net redox states. The observed heterogeneity is therefore biologically compatible with network remodeling rather than a simple uniform depletion of antioxidant capacity. These processes operate within broader mitochondrial and cellular redox-signaling systems [28,33–36].

### 4.4 Specimen matrix and clinical state as sources of heterogeneity

Erythrocytes depend heavily on the pentose phosphate pathway for NADPH and lack nuclei and mitochondria; erythrocyte GR and G6PD therefore reflect a specialized intracellular redox environment. Plasma or serum enzyme measurements may instead be influenced by cellular release, hemolysis, hepatic or renal function, and pre-analytical processing, whereas whole-blood assays combine contributions from multiple cell populations. Matrix-specific interpretation is consequently essential. Persistently high heterogeneity after matrix stratification further suggests contributions from disease severity, treatment exposure, nutritional status, metabolic comorbidity, and assay platform. The treatment-associated differences reported by Klugman et al. illustrate how medication use may reshape peripheral antioxidant enzyme profiles [7].

### 4.5 MCI and potential clinical relevance

Evidence at the MCI stage was particularly sparse. The two erythrocyte GR studies produced effects in opposite directions and do not support GR as a stand-alone early marker. MCI is etiologically heterogeneous, and variation in underlying amyloid and tau pathology may dilute disease-specific metabolic signals [19,25]. Studies that define MCI using established AD biomarkers and simultaneously quantify NADPH, NADP□, glutathione redox couples, and pathway enzymes may be more informative than single-enzyme assays. This multianalyte strategy is consistent with broader evidence that glutathione and lipid-peroxidation markers are altered in cognitive impairment but are strongly affected by matrix and methodology [18–20,30,31].

### 4.6 Research implications

The principal contribution of this review is to define what the current evidence can—and cannot—measure. The next phase of research should move beyond isolated surrogate enzymes toward direct metabolite quantification and integrated pathway phenotyping. Prospective studies should standardize collection time, anticoagulant, centrifugation, hemolysis assessment, and storage; use validated targeted mass-spectrometric or enzymatic methods; and report absolute NADPH and NADP□ concentrations together with their ratio and major related enzyme activities. Longitudinal designs could then determine whether these measures track cognitive decline, established AD biomarkers, or treatment response. Lessons from the implementation of blood-based amyloid and phosphorylated-tau biomarkers emphasize the importance of analytical validation, pre-analytical harmonization, and clinically defined use contexts [24–26].

### 4.7 Strengths and limitations

This review explicitly separated direct NADP(H) measurements from pathway-related enzyme markers and stratified syntheses by disease stage, matrix, and analyte. REML random-effects models, modified Hartung–Knapp intervals, and sensitivity analyses were used to accommodate small and heterogeneous evidence sets. Limitations include few poolable studies, nonuniform assays and units, graph-only outcomes, and an internally inconsistent report. A formal risk-of-bias stratification was not performed; the results are therefore most appropriate for defining the direction of evidence and the principal research gap, rather than for establishing diagnostic thresholds.

## 5. Conclusions

Peripheral NADP(H)-related enzymes may undergo disease-associated remodeling in AD and MCI, but the available literature does not demonstrate a stable and consistent alteration in GR or G6PD. More importantly, direct measurements of NADPH, NADP□, and the NADPH/NADP□ ratio are absent. Standardized, biomarker-defined, and longitudinal studies that directly interrogate NADP(H) metabolism are needed before this pathway can be evaluated for disease stratification, progression monitoring, or treatment assessment.

## Data Availability

All data produced in the present work are contained in the manuscript

## Declarations

### Ethics approval and consent to participate

This systematic review and meta-analysis used published aggregate data and involved no new recruitment of human participants or identifiable individual-level information; additional ethics approval and informed consent were therefore not required.

### Systematic review registration

PROSPERO registration number: CRD420261566620.

### Data availability

All data analyzed in this review were derived from published reports. The curated study-characteristic dataset, effect-size dataset, sensitivity-analysis outputs, and R code are available from the corresponding author on reasonable request. Following acceptance, the authors intend to deposit the reproducible dataset and code in an open repository and add the permanent link to the final version.

### Author contributions

Dongsheng Zhong contributed to the study conception and design, literature search, study screening, data extraction, statistical analysis, and drafting of the manuscript. Dongquan He contributed to study screening, data verification, interpretation of the results, and revision of the manuscript. Jia Chen provided methodological guidance, verified the statistical results, and critically revised the manuscript for important intellectual content. Chenkai Yanyang supervised and administered the study, contributed to the interpretation of the results, and approved the final version of the manuscript. All authors reviewed and approved the final manuscript and agreed to be accountable for the accuracy and integrity of the work.

## Acknowledgments

None

